# For a structured response to the psychosocial consequences of the restrictive measures imposed by the global COVID-19 health pandemic: The MAVIPAN longitudinal prospective cohort study protocol

**DOI:** 10.1101/2020.11.10.20227397

**Authors:** Annie LeBlanc, Marie Baron, Patrick Blouin, George Tarabulsy, François Routhier, Catherine Mercier, Jean-Pierre Després, Marc Hébert, Yves De Koninck, Caroline Cellard, Delphine Collin-Vézina, Nancy Côté, Marie-Pier Déry, Émilie Dionne, Richard Fleet, Marie-Hélène Gagné, Maripier Isabelle, Lily Lessard, Matthew Menear, Chantal Mérette, Marie-Christine Ouellet, Marc-André Roy, Marie-Christine Saint-Jacques, Claudia Savard, on behalf of the MAVIPAN Research Collaboration

## Abstract

**Background:** The COVID-19 pandemic and the isolation measures taken to control it has caused important disruptions in economies and labour markets, changed the way we work and socialize, forced schools to close and healthcare and social services to reorganize in order to redirect resources on the pandemic response. This unprecedented crisis forces individuals to make considerable efforts to adapt and can have serious psychological and social consequences that are likely to persist once the pandemic has been contained and restrictive measures lifted. These impacts will be significant for vulnerable individuals and will most likely exacerbate existing social and gender health and social inequalities. This crisis also puts a toll on the capacity of our healthcare and social services structures to provide timely and adequate care. In order to minimize these consequences, there is an urgent need for high-quality, real-time information on the psychosocial impacts of the pandemic. The MAVIPAN (*Ma vie et la pandémie/My life with the pandemic*) study aims to document how individuals, families, healthcare workers, and health organisations that provide services are affected by the pandemic and how they adapt.

**Methods:** The MAVIPAN study is a 5-year longitudinal prospective cohort study that was launched on April 29^th^, 2020 in the province of Quebec which, at that time, was the epicenter of the pandemic in Canada. Quantitative data is collected through online questionnaires approximately 5 times a year depending on the pandemic evolution. Questionnaires include measures of health, social, behavioral and individual determinants as well as psychosocial impacts. Qualitative data will be collected with individual and group interviews that seek to deepen our understanding of coping strategies.

**Discussion:** The MAVIPAN study will support the healthcare and social services system response by providing the evidence base needed to identify those who are most affected by the pandemic and by guiding public health authorities’ decision making regarding intervention and resource allocation to mitigate these impacts. It is also a unique opportunity to advance our knowledge on coping mechanisms and adjustment strategies.

**Trial registration:** NCT04575571 (retrospectively registered)

## BACKGROUND

The health crisis imposed by COVID-19 is forcing major worldwide social reorganization that will have profound consequences on our society [1]. Affected countries have been attempting to contain the spread of the virus by requiring extraordinary isolation efforts from their populations [2, 3]. One-third of the world’s population (∼3 billion individuals) has, is or will again experience some kind of isolation measures, causing an unprecedented and rapidly evolving psychosocial crisis [4-7]. While biomedical research is relentlessly pursuing its efforts to understand the impact of the disease on infected individuals and to develop new treatments and vaccines, its psychosocial consequences that could permanently affect our wellbeing, the state of our health system, and our society cannot be ignored [7-10].

Failure to address psychosocial and health issues will prolong the impact of the pandemic for years to come. The psychosocial consequences of this health crisis will spare no one, particularly vulnerable individuals, and will persist long after restriction measures are lifted and the pandemic is over [7, 9, 11, 12]. The combination of professional changes, the state of being “at risk”, the possible loss of employment, the resulting economic difficulties, changes in couple and family dynamics, school closures and the reduction of services in health and social services network may all have an impact on the adjustment and development of individuals of all ages [13-17]. This impact will be significant for individuals facing unique contexts or challenges (e.g., older adults, individuals living with a disability, individuals with a chronic or mental health condition, underprivileged families) and will most likely exacerbate existing social, racial and gender inequalities in health and human development [18-25].

The scale of the current COVID-19 mobilization has destabilized several aspects of our health and social services structures. Services are being suspended, others are being maintained or intensified, and new intervention strategies are rapidly being adopted to adjust to containment measures or risk of virus transmission [26, 27]. Service interruptions, amongst others, have an impact on the physical and mental health and subsequent development of already vulnerable individuals [28]. Health and social services workers are experiencing major changes in their practice during this crisis [9, 29]. Many of these workers bear constant witness to the human toll of the pandemic and, all too often, become part of it [9, 30, 31]. This occurs in a context where these workers are subject to the same measures as the rest of the population, thus placing greater demands on their ability to adapt [30, 32, 33]. It is crucial to document practice changes and adjustments of these individuals, who must remain available for their family, colleagues and the population.

Recovering from the pandemic will require a social and economic response that is just as important as the current efforts to minimize the spread of infection [1, 34]. There is an urgent need for information on the evolution of the psychosocial dimensions of health and coping strategies used by our population and our health and social services structures. By comprehensively documenting such information, stakeholders will be in a better position to make timely informed decisions and implement strategies to minimize the expected consequences of the crisis on our mental health and well-being.

The MAVIPAN (*Ma vie et la pandémie*) cohort was developed in response to these individual and collective needs. It was born out of an unprecedented collective effort between the 4 research centers of the Quebec Integrated University Health & Social Services Center and their province-wide academic, governmental, institutional and community partners.

## METHODS

### Aims

Overall, MAVIPAN aims to accelerate the availability of high-quality, real-time evidence within health and social services structures to address, support and minimize ongoing and future, direct and indirect, psychosocial consequences of the COVID-19 pandemic. Working toward that goal, through constantly evolving research questions responsive to the pandemic evolution and knowledge users (KUs) needs, we will document, monitor, and evaluate the following:

i. Individual and family adjustment and mitigation strategies, especially for those considered vulnerable and in high-risk contexts (e.g., What are the psychosocial and professional characteristics of the most vulnerable participants? What are the characteristics of those who seem to be coping well and who may have even improved during confinement? How are families coping over time?);
ii. Healthcare, social services and frontline worker adjustment and mitigation strategies (e.g., What is the role of coping and adaptive strategies on the wellbeing and psychological health of our workers? Are there specific sectors of activity or levels of responsibility that are more vulnerable to adjustment issues? What are the predictive factors of burnout amongst healthcare workers?);
iii. The organization of service structures (e.g., What are the mental health and social service needs non-related to COVID-19 that are not covered or poorly covered by current services? How have some services reorganized to provide appropriate levels of care and minimize barriers to care delivery?) and
iv. The social and economic response (e.g., What are the economic, social and community-based initiatives that have contributed to mental health wellness? What health or social services should be prioritized upon another confinement?).

We have established strategic research priorities under key themes to address our objectives. We have identified Health inequities and mental health as cross-cutting themes across our objectives. Additional key themes have emerged from sources most likely to increase vulnerability during this health crisis: social environment and health, chronic diseases and disabilities, and frontline, health and social workers. Together, these strategic research priorities will be used as an evolving roadmap to assess the level and extent to which we are addressing our research objectives in a way that meets the needs of KUs and state of current knowledge.

### Conceptual Underpinnings

The proposed approach draws from discussions with and lessons learned from KUs and field experts, literature reviews on the psychosocial impacts of disasters, quarantine, and long-term inequalities resulting from crises, and considerations of the strongest study design with the least risk of bias, while considering the complexities of the current and evolving pandemic situation. MAVIPAN is grounded into the (i) Integrated Knowledge Translation (iKT) approach that actively involves KUs throughout the entire research process and its governance to enhance the relevance and uptake of results, [35] and the complementary (ii) SPOR Patient Engagement Framework to foster a climate in which researchers and KUs understand the value of patient involvement [36]. The design, measures, and analyses are further informed by the (i) Model of Psychosocial Impact of Natural Disasters that specifically addresses coping mechanisms and mitigation strategies during traumatic events, [37] and the (ii) Dahlgren and Whitehead’s Model of the Social Determinants of Health that identifies the environmental, social, and individual spheres of influence that hinder or enhance the health of individuals and create inequities between populations [38]. We will add to this model by considering structural and political determinants of health, which are emerging in the critical race literature [39-41].

### Study Design

MAVIPAN is a mixed-methods based, prospective, observational, longitudinal cohort where participants will be followed over a 5-yr period [42]. We will collect quantitative and qualitative data at time of recruitment and then according to the 4 expected phases of the pandemic evolution: the impact phase we are experiencing, the turning point phase when the crisis is brought under control, the recovery phase, and the post crisis following a new “normal”, accounting for additional infection waves and major events (e.g., vaccines) (Figure 1). The longitudinal aspect of the cohort sets itself apart. The collection of information related to the same individual at multiple points throughout the evolution of the crisis allows for a unique understanding and insights into mechanisms at play, temporal relationships with key crisis events, and the persistent or transient nature of the psychosocial impacts, and can inform when and how to intervene [43, 44]. The use of mixed methods is well-suited for this proposal and adds to its significance [45]. Quantitative data measure indicators, determinants, and impacts (short, mid and long term). Qualitative data build a reflexive approach into the determinants, and will be central to exploring and identifying unexpected impacts and adaptation strategies experienced by participants.

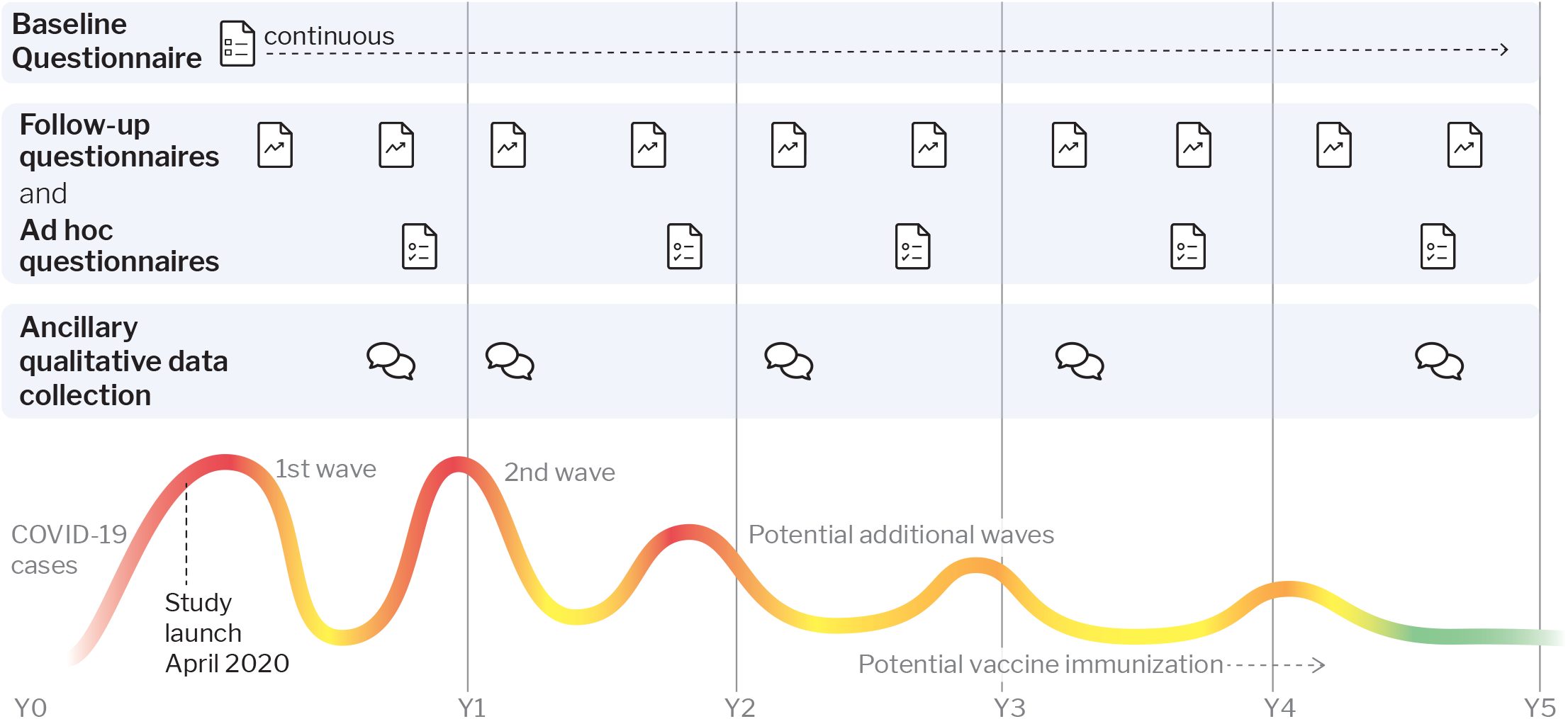

### Participants

MAVIPAN is open to any individual aged 14 and over who understands French or English across the Province of Quebec, the epicentre of Canada’s COVID-19 epidemic [46]. Within this province, we have been and will continue to reach individuals in urban, suburban and rural areas where different numbers of COVID-19 cases (from no cases to hotspots) are found. We are particularly invested in recruiting vulnerable populations (e.g., older adults, individuals living with a disability or a chronic/mental health condition, minorities, child protection families, individuals living in institutional settings) and populations that have become vulnerable because of the COVID-19 context (e.g., healthcare and social services workers, adolescents and young adults, caregivers).

### Recruitment

We continue to systematically recruit across the province, through our website [47], lead media, social media and networks (e.g., Twitter, Facebook), and mass diffusion across healthcare establishments, universities and large networks. We are supported by regional Public Health Directions from healthcare establishments across the province. We have established and continue to seek collaborations with urban and rural cities (e.g., City Halls) who promote MAVIPAN through their networks. We developed a recruitment plan tailored to our vulnerable populations, that includes collaborations with (i) key clinical departments and programs (e.g., COVID-19 clinic) and communication offices within healthcare establishments across the province to directly reach patients and clients, (ii) community-based organizations and (iii) provincial thematic networks or associations.

### Retention Plan

We recognize the challenge of loss to follow-up in prospective cohorts [43, 44, 48]. We have developed a retention plan that includes, but is not limited to: study branding and publicity, incentives (e.g., annual gift certificates), personalized email messages, intermittent lay language summary of findings disseminated to participants, and an individualized study page to keep participants informed and engaged [49-51].

### Quantitative Data Collection

We collect quantitative data through online questionnaires using the REDCap electronic data acquisition platform that is maintained by Université Laval Collaborative platform for large-scale and sustainable data collection Pulsar [52]. Registration, consent and questionnaires can be completed on different digital devices, in French or English. We also provide support for people who do not have access to the Internet or to a computer, have limited digital literacy, or have a disability (e.g., manual-gestural language, research assistant).

Participants can register at any point over the course of the study and complete a thorough baseline questionnaire (30-45 min). Additional questionnaires, up to 4 per year, will be tailored to key events in the crisis evolution (e.g., second wave) and change in restrictive measures (e.g., closing of schools). These include a brief (15 min), standardized follow-up questionnaire which we intend to administer at least twice a year and ad hoc questionnaires (<30 min) aligned with key events (e.g., vaccine). A notice and then one reminder is sent to participants when a new questionnaire becomes available. Participants are given a week to complete questionnaires (i.e. they can start filling a questionnaire, stop at any time and come back to their saved questionnaire). Questionnaires have been and will continue to be developed and pilot tested with key experts and KUs, using brief (instead of exhaustive) validated measures when available.

### Measures

We selected well-validated measures based upon our theoretical models, Public Health recommendations, expert consensus, and KUs’ inputs [35, 37, 53]. We document health, social, behavioral and individual determinants and psychosocial impacts of the pandemic for baseline and follow-up questionnaires (Figure 2) [54-70]. We further added specific measures and indicators for vulnerable populations, such as disease management, changes in life circumstances attributable to the pandemic, or caregiver burden. Participants have the opportunity to fill in open-ended questions addressing current or expected challenges, helpful innovations, hopeful moments, and additional topics they would want to see addressed in future questionnaires.

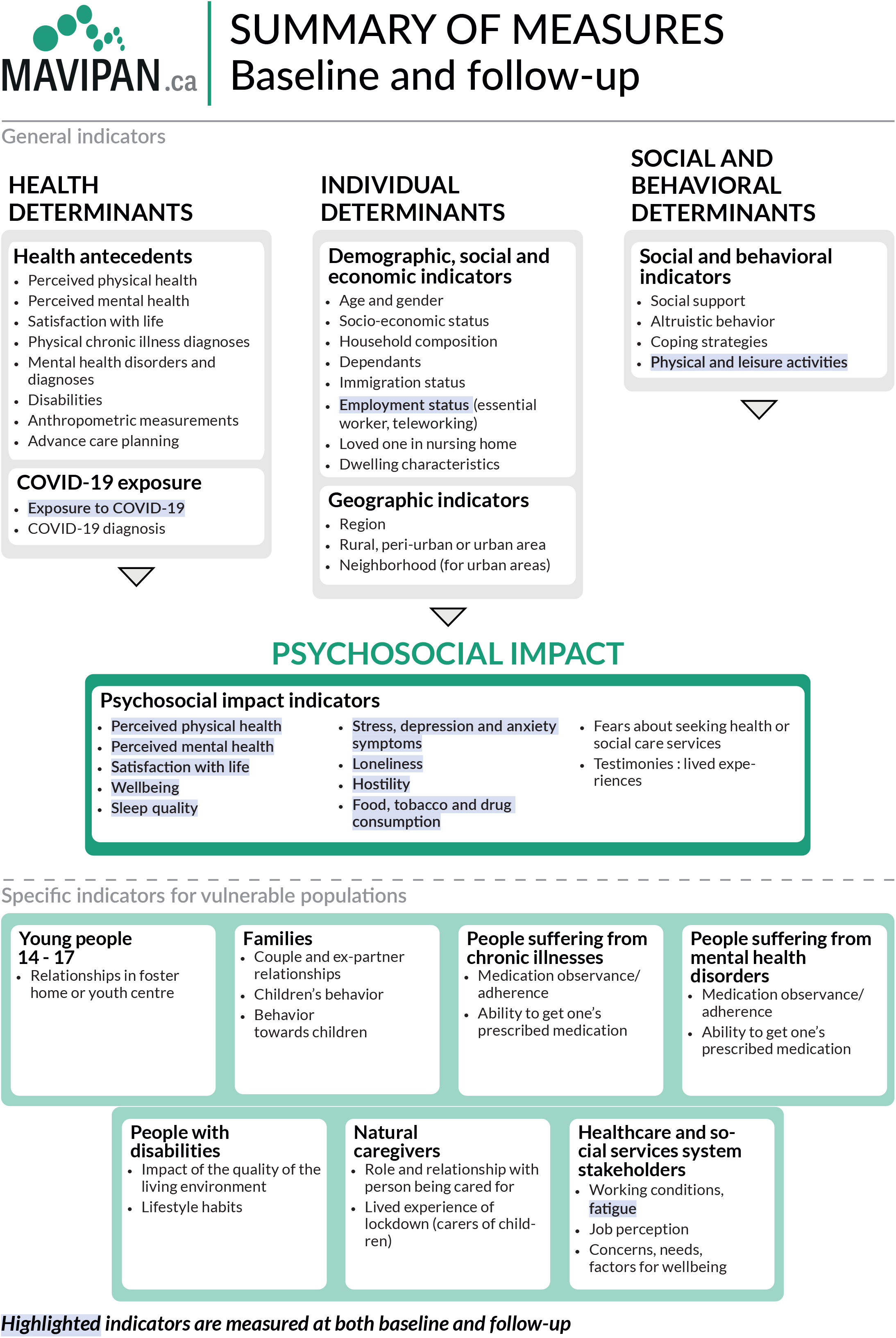

### Linkage

At registration, we ask participants if they agree to be contacted for additional research opportunities. This allows us to add ancillary protocols (e.g., interviews with subsets of participants) in response to the pandemic evolution, our findings, and the needs of KUs, thus substantially improving the quality and relevance of the information that is gathered. Furthermore, this allows for opportunities to link MAVIPAN with provincial, national and international COVID-19 related initiatives, thus fostering dynamic, multidisciplinary collaborations leading to increased impact.

### Qualitative data collection

Each ancillary qualitative protocol will be unique yet (i) will share common elements of their interview guide (e.g., mitigation strategies, impact of the pandemic) and (ii) rely on best practices for the conduct of its activities [71, 72]. We will conduct semi-structured interviews and focus groups mainly through securitized online medium (e.g., Zoom, Microsoft Teams) for the time being and will adapt as the restrictive measures are lifted. Length and number of participants will be tailored to each research question. Additional approaches (e.g., observations) could be added if relevant.

### Ethics

We have worked and continue to work in close collaboration with our Research Ethics Committee, who has been instrumental in designing this “living” cohort. We have set templates and procedures in place allowing for an agile process and rapid response (e.g., within days) to new questionnaires and ancillary protocols being submitted. All study procedures have been approved by the respective Research Ethics Committee of all participating institutions.

### Data management

We recognize that longitudinal studies require an appropriate data infrastructure that is sufficiently robust to withstand the test of time [43, 44, 48]. MAVIPAN operates using the REDCap system, a HIPAA compliant secure data entry system, housed within Université Laval’s Pulsar infrastructure. Data management is under the shared responsibility between the research team and Pulsar’s highly qualified personal. This setting ensures the highest of standards (i.e. standardized data collection procedure, secured data storage, quality control, daily back-up system) in a sustainable infrastructure that guaranties housing of the data for years to come.

### Sample Size

We propose a cost-effective, time sensitive, non-probabilistic purposive sampling paired with online sampling and a snowballing technique without size restriction. Based of recruitment rate so far (2,800 participants in the first 5 months of the study), we expect to recruit 5,000 participants by the end of 2021 and 7,500 participants by the end of the study. We recognize bias and limitations associated with this approach (e.g., sampling error, self-selection, lack of representation of population) [48]. We have included key sociodemographic questions that will enable us to compare and weight data to provincial and national standards. We have used similar validated questions as key institutions such as Statistics Canada to further ensure comparability of our findings.

### Analysis

We will pursuit analysis under a mixed-method umbrella, with both sequential and simultaneous analysis of quantitative and qualitative data, to strengthen the breadth and depth in our capacity to answer our research questions [71]. Findings from the quantitative analysis will inform phases of qualitative data collection and hypotheses derived from qualitative analysis will inform subsequent quantitative component. Triangulation will be used to corroborate our findings and help explain paradoxes or inconsistencies emerging within the qualitative or the quantitative analysis.

Statistical analyses will involve both cross-sectional and longitudinal methods and will be of two-folds. First, in a cross-sectional fashion, we provide constant, descriptive information for KUs, enabling them to understand the characteristics of those individuals who are faring well and not so well during the present crisis [73]. In doing so, we help KUs identify high-risk individuals and families as a function of different sociodemographic characteristics (e.g., sociodemographic, occupational) as they relate to mental health problems, social and health behaviors, identified needs and the use of health and social services (i.e. health inequalities). These analyses will help identify populations that may be easily targeted for immediate health services or intervention to improve the state of our service structures.

Then, we will conduct analyses that will help us identify empirically-derived groups in adjustment as a function of time or other variables (i.e. identify individual differences in risk with the added benefits of multiple measures of adjustment). We will conduct generalized linear mixed models that will allow us to make associations between events that unfold and characterize the current crisis and individual adjustment across time, informing resilience trajectories, coping and adaptation mechanisms as well as cumulative burden experienced by subgroups of the population [74]. In-depth analyses of the factors contributing to the health and well-being (or lack of) of these subgroups will further inform on the mechanism underlying the aggravation of health inequalities.

Lastly, we will develop analytical strategies tailored to each research question. These strategies will likely include range of methods appropriate for cross-sectional, longitudinal, linked data and causal modelling when relevant, adjusting for missing data where necessary. We will account for sex and gender-based analysis and use an intersectionality approach to explain potential comparisons with emerging key factors (sex, age, and race) in outcomes of COVID-19. We perform analysis using SAS (version 9.2) or R (version 4.0.1) software package.

Our overall qualitative approach will rely on thematic analysis (although other approaches, such as a narrative approach to qualitative inquiry whereby accounts of experience are explored from the life perspective, could be added if relevant) [71, 75]. Audio or video recorded interviews will be transcribed in verbatim, de-identified, and verified against actual recordings by team members. We will audio or video record focus groups, which will be complemented by the moderator and an observer observational notes. We will follow best practices for data management and organisation, coding, and analysis, using the most relevant software (NVivo, QDA Miner or Noldus Observer) [71, 75].

Furthermore, throughout the study, we will conduct cross-comparisons between the ancillary protocols (including a meta-analysis of all qualitative results), which will contribute to a conceptual framework on the health impact and adaptation strategies during a world-wide pandemic.

### Transparency of Research & Data Sharing

Transparency of MAVIPAN will be evident in the clarity and completeness of datasets, codebooks and supporting documentation, many already available [47]. The substantial investments necessary to build these large studies and the unprecedented nature of this health crisis argue for optimal utilization of MAVIPAN. Data produced as a result of this study will be shared in line with the Canadian Institute of Health Research joint statement on sharing research data and findings relevant to this coronavirus outbreak. Resulting publications will be open access.

Researchers and collaborators will be able to submit research questions and obtain access to data sets. Questions being investigated will be posted on our website to avoid redundancy and promote collaboration within the research community, healthcare institutions, public health agencies, government officials and community organizations.

### Governance

Large longitudinal cohort studies are demanding and require sound and sustainable infrastructure and governance. We have set in place an equitable, inclusive and sustainable Governance Plan that fully includes citizens, patients, other KUs, experts, and representatives from participating research centers, health establishments, and organizations across the Province.

We have established (i) a Steering Committee (quarterly meetings) that provides strategic leadership, including research question priorities, milestones and national and international collaborations, facilitate research and knowledge translation activities, (ii) an Executive Committee (monthly meetings) that reviews requests for collaboration and submission of research questions (i.e. alignment with research priorities and feasibility), progress and challenges of ongoing work, approval of publications to be submitted, and scholarship process for graduate students, and (iii) a Lead Research Team (bi-weekly meetings) that will handle the day-to-day operations of MAVIPAN.

### Study Status

MAVIPAN was developed and launched within six weeks of the first confinement in March 2020. It was designed with the aim to be flexible and adapt according to the pandemic evolution and resolution and its associated restrictive measures in the upcoming 5 years. The study is currently opened for enrollment.

### Knowledge translation plans

We have a well-defined iKT approach where KUs are involved throughout the research process and contribute to just-in-time diffusion and dissemination of research progress and outputs. We are providing, on an as-needed basis, following the crisis evolution, personalized (i.e. as a function of region or clientele) updates to KUs. Our KUs and collaborators are helping build community partnerships and assisting us with translation and dissemination of findings. Our bilingual website [47] and those of our collaborators will be an important tool for communicating our findings to other populations, stakeholders and research groups in Quebec, Canada and internationally. We will have plain language summaries posted on the website. Moreover, as the launch of the cohort drew media attention, we will build upon this interest, disseminating findings through news media and social networks. Furthermore, each of the research centres and healthcare institutions involved engages actively in KT towards practitioners and professionals, stakeholders and administrators, service users and other sectors of the population. Each of these platforms will be leveraged to ensure that pertinent information is constantly transmitted.

## DISCUSSION

### Anticipated Outcomes and Impact

The COVID-19 health crisis has caused an unprecedented scientific collaboration. Worldwide, scientists of different countries and background have come together, rapidly sharing the most recent and relevant information about the pandemic. MAVIPAN takes part in this international scientific collaboration. As the epicentre of the pandemic in Canada, the province of Quebec is now a unique living laboratory to measure, understand and act on the impact of public health measures on population’s health and well-being. Results produced with MAVIPAN add new, unique and most relevant information to other governments and population in Canada, North America and worldwide to adjust the public health response in the next months and years. As other important waves of virus outbreaks are expected to take place, the MAVIPAN experience is central to improve the public health response.

MAVIPAN has the potential to be a critical component of the response to COVID-19 as it can initiate new rapid response to unmet needs. It can support institutions in the mental health and social services network and inform the evidence base underpinning the deployment and organization of services in times of crisis and in the recovery period. MAVIPAN’s unique infrastructure will increase the potential for data collection to be harmonized, shared and integrated across COVID-19 related initiatives. It will promote an agile, multidisciplinary and collaborative approach to research and address challenging and important COVID-19 research questions in a concerted and high-impact manner. Findings from MAVIPAN will improve our understanding of the psychosocial impacts, the coping mechanisms and adaptive strategies that have emerged from the restrictive measures of this unprecedented pandemic.

### Limitations, anticipated problems and solutions

A major source of potential bias in cohort studies is due to losses to follow-up [44]. Cohort members may migrate or refuse to continue to participate in the study. We have put a sound and proven-effective retention plan in place. Open registration throughout the study compensates, to some extent, for loss to follow-up. We will also assess the seriousness of the bias in the measures of effect of exposure and outcome that this may case in incorporating this issue in our analysis plans. We have overcome issues of variations in data collection that sometime occurs in multi-centered studies. All quantitative data collection is done through a unique platform and we have established a common template for all qualitative activities. The collaboration with Pulsar ensures that issues of management of such a large database (i.e. cost) are minimized and sustainability of the database secured.

## CONCLUSION

Launched in April 2020 across the Province of Quebec (Canada), MAVIPAN documents health, mental health, social, behavioral, environmental and individual determinants and psychosocial impacts of the pandemic. It is a unique initiative that will contribute in the upcoming months and years to a coherent and integrated mitigation strategies response from health, mental health and social services workers, researchers, public health authorities, policymakers, and the healthcare system, within and across jurisdictions in Canada.

## Data Availability

N/A

## DECLARATIONS

### Ethics Approval

This study is approved by the Ethics Committee of the Primary Care and Population Health Research Sector of the CIUSSS de la Capitale-Nationale (Reference number: 2021-2043).

### Competing Interests

The authors declare that they have no competing interests.

### Funding

Funding for this study comes from discretionary funds from the four Research Centers of the Quebec Integrated University Health and Social Services Center (Vitam: Centre de recherche en santé durable, Centre de recherche universitaire sur les jeunes et les familles (CRUJeF), Centre interdisciplinaire de recherche en réadaptation et intégration sociale (CIRRIS), Centre de recherche CERVO).

### Author’s contribution

The following authors conceived the study, co-wrote the first draft, and made critical revisions to the manuscript: (AL, MB, PB, JPD, YDK, CM, GT, CM, FR). The following authors participated to the design of the study and made critical revisions to the manuscript: (CC, DCV, NC, ED, RF, MHG, MI, LL, MM, CM, MCO, MAR, MCSJ, CS). The following authors contributed to the design of the study and provided revisions of the manuscripts (AB, MP, MPG, DL, MEL, HW, PA, GR, AV, MHM, MG, ESH, AG, EG, DN, JT, GD, MCS, SBB, NB, MFD, PM, LV). All authors approved the final version of this manuscript.

## Acknowledgements

MAVIPAN Research Collaborative would like to acknowledge the following research team members for their time and implication in this research: Léa Langlois, Frédéric Cantin, Josiane Lettre and Geneviève Picher. We would also like to acknowledge the ongoing support and collaboration of the PULSAR team, including Laurence Dionne-Bibaud, Audrey St-Laurent, Marie-Andrée Lévesque and Carole Artault, the CIUSSS-CN Ethics committee, the CIUSSS de la Capitale-Nationale, the CISSS Chaudière-Appalaches, CISSS Bas-Saint-Laurent, CISSS Côte-Nord, and all our partners in the project. For the complete list, please go to www.mapivan.ca. Finally, we would like to give special thanks to all citizens and participants who give us feedback and support in the MAVIPAN project.

## Notes

### Competing Interest Statement

The authors have declared no competing interest.

### Clinical Trial

NCT04575571. This study was registered retrospectively. Considering the very short timeframe and the urgency surrounding the pandemic as well as the fact that registering observational studies is not mandatory, the priority was given to the implementation of this project before registering it.

### Summary of Updates

Missing author was added in the authorship list.

